# Updating urinary microbiome analyses to enhance biologic interpretation

**DOI:** 10.1101/2021.09.30.21264391

**Authors:** Nazema Y. Siddiqui, Li Ma, Linda Brubaker, Jialiang Mao, Carter Hoffman, Lisa Karstens

## Abstract

**Objective:** An approach for assessing the urinary microbiome is 16S rRNA gene sequencing, where a segment of the bacterial genome is amplified and sequenced. Methods used to analyze these data are rapidly evolving, although the research implications are not known. This re-analysis of an existing dataset aimed to determine the impact of updated bioinformatic and statistical techniques.

**Methods:** A prior Pelvic Floor Disorders Network (PFDN) study compared the urinary microbiome in 123 women with mixed urinary incontinence (MUI) and 84 controls. We used the PFDN’s unprocessed sequencing data of V1-V3 and V4-V6 16S variable regions, processed operational taxonomic unit (OTU) tables, and de-identified clinical data. We processed sequencing data with an updated bioinformatic pipeline, which used DADA2 to generate amplicon sequence variant (ASV) tables. Taxa from ASV tables were compared to OTU tables generated from the original processing; taxa from different variable regions (e.g., V1-V3 versus V4-V6) after updated processing were also compared. After updated processing, data were analyzed with multiple filtering thresholds. Several techniques were tested to cluster samples into microbial communities. Multivariable regression was used to test for associations between microbial communities and MUI, while controlling for potentially confounding variables.

**Results:** Of taxa identified through updated bioinformatic processing, only 40% were identified originally, though taxa identified through both methods represented >99% of sequencing data in terms of relative abundance. When different 16S rRNA gene regions were sequenced from the same samples, there were differences noted in recovered taxa. When the original clustering methods were applied to reprocessed sequencing data, we confirmed differences in microbial communities associated with MUI. However, when samples were clustered with a different methodology, microbial communities were no longer associated with MUI.

**Conclusions:** Updated bioinformatic processing techniques recover many different taxa compared to prior techniques, though most of these differences exist in low abundance taxa that occupy a small proportion of the overall microbiome. Detection of high abundance taxa are not significantly impacted by bioinformatic strategy. However, there are different biases for less abundant taxa; these differences as well as downstream clustering methodology and filtering thresholds may affect interpretation of overall results.

## Introduction

The urinary microbiome is being investigated in multiple bladder conditions. There are now several reports demonstrating differences in urinary microbiota in women with recurrent UTI (1, 2), urgency urinary incontinence (3, 4), and mixed urinary incontinence (5) when compared to matched controls without these symptoms. In most of these studies, 16S rRNA gene sequencing has been employed as a culture-independent method of detecting urinary bacteria. When using sequencing to detect bacteria, DNA is extracted from a biological sample, polymerase chain reaction (PCR) is used to amplify and sequence segments of the 16S rRNA gene, and bioinformatic tools are used to match the recovered sequences with those existing in a reference database. Results are reported as taxonomic groupings (i.e., taxa). These steps allow investigators to identify the bacterial taxa contained in a sample. Next, the recovered taxa can be compared between participant cohorts using statistical analyses to discern if there are differences between phenotypic groups.

The bioinformatic steps outlined above depend on multiple computational components, which have been rapidly evolving. Many prior analyses were performed with reference databases that have not been recently updated, such as Greengenes^1^. Furthermore, the Greengenes database does not have substantial representation of urinary bacteria and thus may not be the optimal reference database for identification of microbiota within a urine sample.(6) Regardless of the reference database that is selected, bioinformatic workflows rely on algorithms that group raw sequencing data based on similarities. These algorithms are rapidly evolving, and when updated or refined, could potentially alter bacterial identification results. Previously, researchers would group raw sequences into operational taxonomic units (OTUs) based on similarity, then compared these OTUs against reference databases to identify the bacterial taxa. Many now advocate for grouping raw sequences using amplicon sequence variant (ASV)-based methods, where sequences are grouped based on their exact nucleotides. In ASV-based methods, the error score assessing the confidence of sequencing results at each nucleotide is incorporated such that algorithms can better detect true biologic sequences versus those generated by sequencing error. Furthermore, ASV-based methods have the ability to identify bacterial taxa at finer resolution (e.g., genus and species levels where previously identifications were at higher taxonomic levels such as the family level). Studies that were performed prior to these updates in bioinformatic workflows may benefit from re-analysis.

Separate from bioinformatic components of analyses, the statistical methods used to analyze microbial data are also evolving. Prior studies have used methods such as Dirichlet Multinomial Mixture (DMM) modeling (7), which adopt simplistic distributional assumptions on the microbiome compositions, and linear discriminant analysis effect size (LeFSE) analysis^2^, which is a nonparametric cross-sample test that utilizes linear discriminant analysis (LDA) to construct test statistics assisted by classical univariate tests for feature selection. Neither approach adequately accounts for all of the key characteristics of microbiome data such as their compositional constraints, complex cross-sample heterogeneity, and sparse counts of certain taxa. Thus, high dimensional microbial datasets that are used in these types of compositional or community-based analyses fail to meet the underlying assumptions that are needed for the statistical techniques. Rather, more recently developed tree-based models(8, 9), community-based analyses(10), or other models that more truthfully account for the distributional characteristics may be needed, especially in view of the limited sample sizes in most studies. These modeling techniques are currently under further development and may be able to better detect the true signal within a dataset.

For datasets with robust findings, updated analytic techniques should not substantially alter major findings. However, urinary microbiome results could be especially prone to bias or skew from different analytic techniques, since small differences are magnified in low biomass environments. We hypothesized that updated analyses would enhance precision and allow for more clarity with biologic inferences and thus we used updated techniques to re-analyze raw sequencing data generated in a prior study. Our primary objective was to determine whether taxonomic identifications substantially differ with an updated bioinformatic pipeline. Secondary objectives were to compare taxonomic identifications based on the 16S rRNA gene variable region used, and to assess whether tree-based modeling strategies enhance our ability to differentiate microbial community profiles between women with mixed urinary incontinence and controls.

## Methods

After Duke University Institutional Review Board approval (Pro #00102155), we conducted a re-analysis of sequencing data generated from the Human Microbiome Study in the Effects of Surgical Treatment Enhanced with Exercise for Mixed Urinary Incontinence HMS-ESTEEM Study.(5) The HMS-ESTEEM study was a supplemental translational study embedded within the ESTEEM randomized trial (11) conducted by 8 clinical sites within the Pelvic Floor Disorders Network (PFDN).^3^ Briefly, this was a cross-sectional analysis of microbiome data obtained from women with mixed urinary incontinence (MUI) and age-matched controls. The strict inclusion and exclusion criteria for participants (207 women, 123 with MUI and 84 age-matched controls) are detailed in prior publications.(12, 13) Women completed validated questionnaires to assess urinary symptom burden and to confirm appropriate categorization into MUI and control groups. Additional questionnaires were administered to gather data about hormonal therapies, sexual activity, recent infections, and the presence of any vaginal medications. Urine samples were obtained via transurethral catheterization and stored in a DNA protectant (Assay Assure™, Sierra Molecular Corporation, Incline Village, NV, USA). Samples were transferred with cold packs via overnight shipping to a central laboratory where they were stored at -80^○^C. After all samples were collected, DNA was extracted and subjected to polymerase chain reaction (PCR) amplification and 16S rRNA gene sequencing as previously described.(12) For each sample, two separate 16S rRNA gene segments (i.e., the V1-V3 and V4-V6 hypervariable regions) were sequenced (Figure 1).

**Figure 1:**
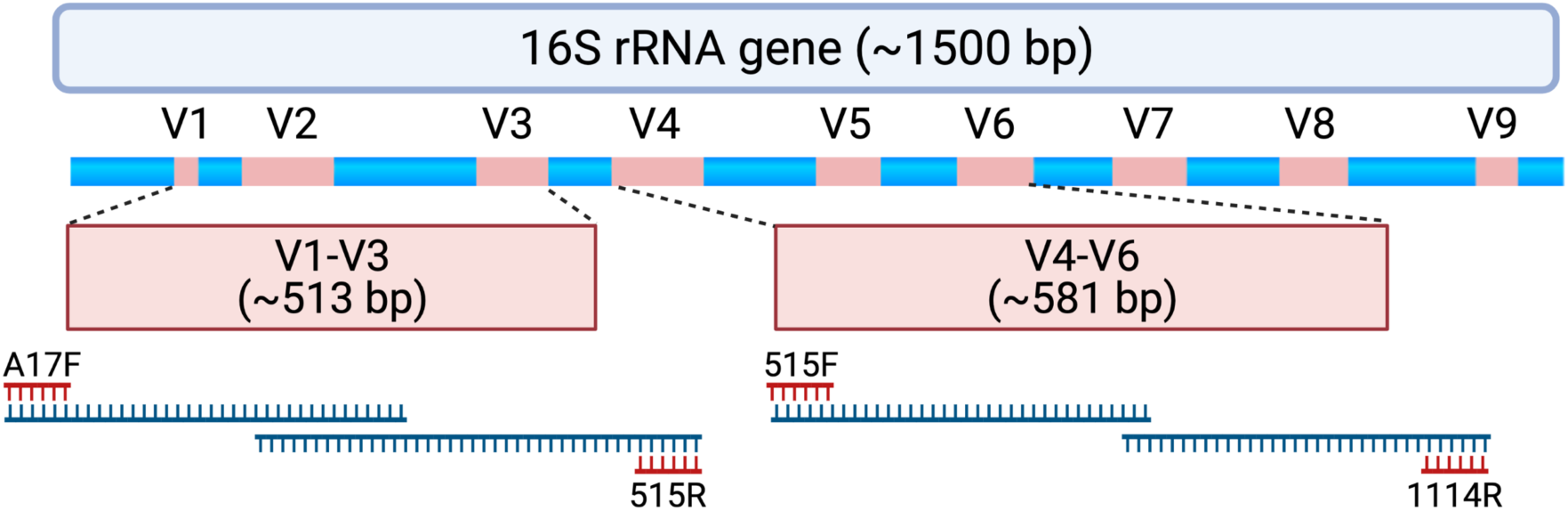
Schematic depicting the 16S rRNA gene and approximate locations of variable regions (V1-V9) that can be selected for amplicon-based sequencing. Samples assessed for this study underwent polymerase chain reaction (PCR) to amplify the V1-V3 region using PCR primers A17F and 515R. When forward and reverse reads are merged, the entire V1-V3 region spans 513 ± 22 base pairs with approximately 87 base pairs of overlapping sequence. Samples also underwent PCR to amplify the V4-V6 region using PCR primers 515F and 1114R. When forward and reverse reads are merged, the entire V4-V6 region spans 581 ± 2 base pairs with approximately 19 base pairs of overlapping sequence. For this study, forward and reverse reads were generated on an Illumina MiSeq platform, which creates sequencing reads of approximately 300 base pairs in length. The initial and final portions of each sequencing read tend to contain lower quality sequence (i.e., lower confidence scores with nucleotide assignment) that could be adjusted or truncated in a DADA2 processing pipeline. As such, paired end reads without a substantial amount of overlapping sequencing may not be able to be merged. Created with BioRender.com.

### Bioinformatics

We obtained unprocessed sequencing files housed under Sequence Read Archive Bioproject #703967 (14), previously generated OTU tables, and associated clinical data from the PFDN data coordinating center. First, we repeated sequence processing using updated techniques. Differences between original and updated processing are illustrated in Figure 2. In the original analysis, sequencing data were processed using the Illumina BaseSpace 16S Metagenomics App version 1.0.1. This software classifies raw sequencing data using ClassifyReads, a high-performance implementation of the Ribosomal Database Project (RDP) classifier (15), and compares classified sequence reads against the Greengenes reference database to identify bacteria. The output is an OTU table, which is a designation of relative proportions of different taxonomic groups that each sample contains. In the updated analysis, raw sequences files were processed using DADA2 (16) (v 1.14.0), then mapped to the SILVA reference database^4^ (v 132) with the RDP classifier. The end result is an ASV table, which is similar to an OTU table while also taking sequencing error into account when grouping sequences together. Data were further processed and visualized in R using phyloseq(17) (v. 1.26.1) and microshades^5^ (v. 0.0.0.9). Taxa identified in the original analysis (i.e., OTU table) and updated analysis (i.e., ASV table) were compared to assess for similarities and dissimilarities based on the sequence processing approach.

**Figure 2:**
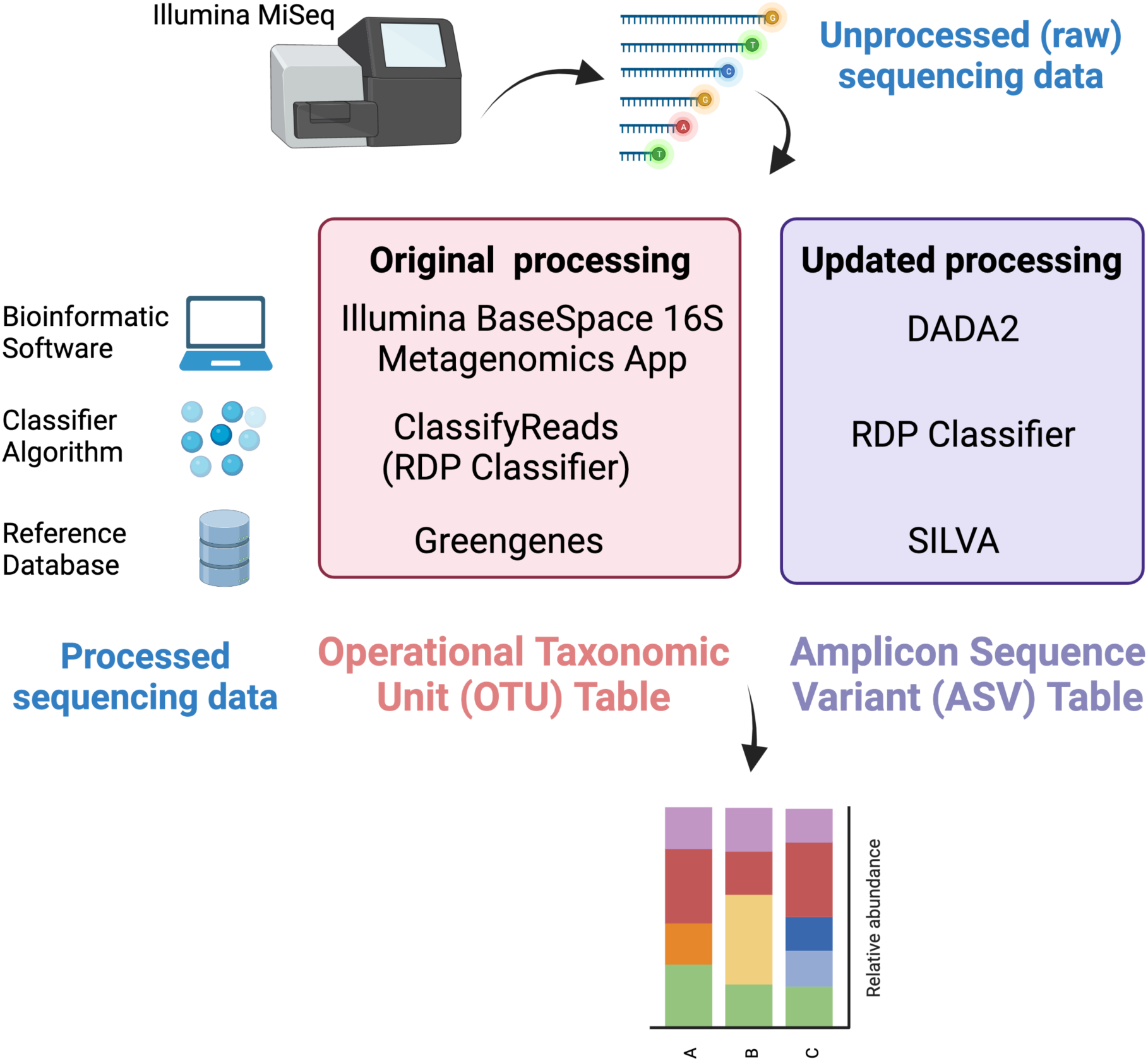
Comparison of original and updated processing. Unprocessed sequencing data generated from an Illumina MiSeq platform were obtained. The figure illustrates differences in the bioinformatic software, classifying algorithm, and reference databases between original and updated processing. The output from original processing was an OTU table. The output from updated processing was an ASV table, which is similar to an OTU table but also adjusts for sequencing errors. Both tabular outputs can be incorporated into Phyloseq objects in R to visualize data as stacked bar plots. Created with BioRender.com.

### Amplicon Comparison

When performing 16S rRNA gene sequencing, generally one or more variable regions (commonly V1-3 or V4-6) of the 16S rRNA gene are amplified and sequenced; the segment of the variable region chosen is referred to as an amplicon. Different amplicons contain distinct regions of DNA, vary in length, and may have different representation in reference databases. As such, one amplicon may identify a specific bacterium at higher resolutions or in a more specific manner than another amplicon targeting a different variable region. Each DNA sample in this study was subjected to sequencing using two different amplicons, one targeting the V1-V3 variable region and the other targeting the V4-V6 variable region. We compared taxa identified from both amplicons for similarities and dissimilarities.

### Rare ASVs: Filtering Thresholds

ASV tables generated from the updated bioinformatic analysis were incorporated into downstream statistical analyses. First, data were filtered such that any taxa below a threshold were considered noise and removed from the dataset. For this dataset, we explored multiple filtering thresholds. Low filtering threshold are most permissive and keep the most data, but increase computational demands, and may lead to spurious results. Higher filtering thresholds remove more data but there is a risk of potentially removing rare taxa that may be important.

### Clustering into Microbial Communities

In trying to infer clinical implications from microbial datasets, samples are often clustered into groups based on those that contain similar bacterial taxa. This is a method of dimensionality reduction where multiple bacterial taxa can be considered together as a microbial community. In the original analysis, taxonomic clustering was performed using Dirichlet Multinomial Mixture (DMM).(7) In this method, the investigator must assign the final number of clusters that are desired. This is achieved by reviewing results with different numbers of clusters and selecting the final number of clusters that qualitatively seems to make sense. We repeated DMM clustering on ASV data from the updated bioinformatic analysis using the same number of clusters that were selected in the original publication. However, since selecting the number of clusters can introduce bias, we also evaluated a nonparametric mixture model called Dirichlet tree multinomial mixture (DTMM)^6^, which automatically adapts the number of clusters based on the complexity of the data. The two methods also differ in the way cross-sample heterogeneity is modeled, resulting in the phenomenon that DMM clusters samples to be highly influenced by the “dominant”, or most abundant taxa in a sample. In contrast, DTMM more effectively incorporates less abundant taxa when creating clusters of similar samples by utilizing the phylogenetic tree to enrich the modeling on cross-sample variability. Whether obtained through DMM or DTMM methods, final clusters are considered microbial communities.

### Testing for Associations between Microbial Communities and MUI versus Control Phenotype

This updated analysis (as well as the original analysis) assessed microbial communities from DMM and DTMM clustering methods in multivariable generalized linear models with a logit link, to see if urinary microbial communities were associated with MUI versus control phenotypes. Both original and updated analyses incorporated covariates though these were chosen differently, as detailed in Supplemental Table 1. Along with microbial community types, the original and updated analyses incorporated age, ethnicity, body mass index (BMI), and smoking status in final multivariable models. In the updated analysis we also included a composite variable that incorporated menopausal and hormonal status as one of three categorical options: 1) pre-menopausal; 2) post-menopausal with any estrogen hormone use (topical, vaginal, transdermal, oral); and 3) post-menopausal without hormone use. In addition, we included vaginal pH, dichotomous history of recurrent UTI, and number of days from the most recent catheterization (calculated based on last prior recorded catheterized urine sample or urodynamic assessment) as covariates in the updated analysis. These variables have been proposed as “desired” within recently published standards for urinary microbiome research.(18) Both the original and updated analyses considered clinical site where samples were acquired, though site was managed differently in original versus updated models (see Supplemental Table 1 & Supplemental Figure 1). Clustering and multivariable modeling were performed in R.

## Results

The unprocessed sequencing data from 207 samples (123 MUI and 84 controls) that were sequenced using 300bp paired-end reads from V1-V3 and V4-V6 variable regions resulted in taxonomic data available from 173 samples for the V1-V3 region and 194 samples for the V4-V6 region after reprocessing the sequencing files. There was significant data loss of approximately 25% of the samples from the V1-V3 region with attempts to merge forward and reverse reads. Sequencing reads from the V4-V6 region, which provides a longer amplicon, were unable to be merged because of lack of enough overlapping sequence (see Figure 1). As such, we used forward reads only for both amplicons in subsequent analyses. Median sequencing depth and the recovered taxa from the ASV table are summarized in Table 1 and compared to those reported in the original analysis, which only reported on the V4-V6 amplicon. When comparing the originally processed (OTU table) and updated (ASV table) V4-V6 data, 329 genera were only identified with original processing, 426 were only identified with updated processing, and 289 were overlapping and identified with both (Figure 3a). Though there were many non-overlapping genera, these were represented in the small proportion of the low abundance sequences from all samples (Figure 3b).

**Table 1:** Sequencing data & Recovered taxa

|  | Median (range)<br>sequencing depth | # Phyla | # Classes | # Orders | #Families | #Genera |
| --- | --- | --- | --- | --- | --- | --- |
| V1-V3* | 24,862<br>(1,021 – 670,442) | 29 | 63 | 143 | 220 | 545 |
| V4-V6* | 29,105<br>(5,029 – 187,593) | 27 | 73 | 182 | 256 | 721 |
| Original<br>analysis<br>V4-V6 (5) | 80,949<br>(12,517 – 450,993) | 28 | 60 | 82 | 191 | 581 |

\* Due to technical issues when merging forward and reverse reads while using a pipeline that generates amplicon sequence variants (ASVs), only forward reads were used in repeat bioinformatic processing. Both forward and reverse reads were used in originally processed and reported data.

**Figure 3:**
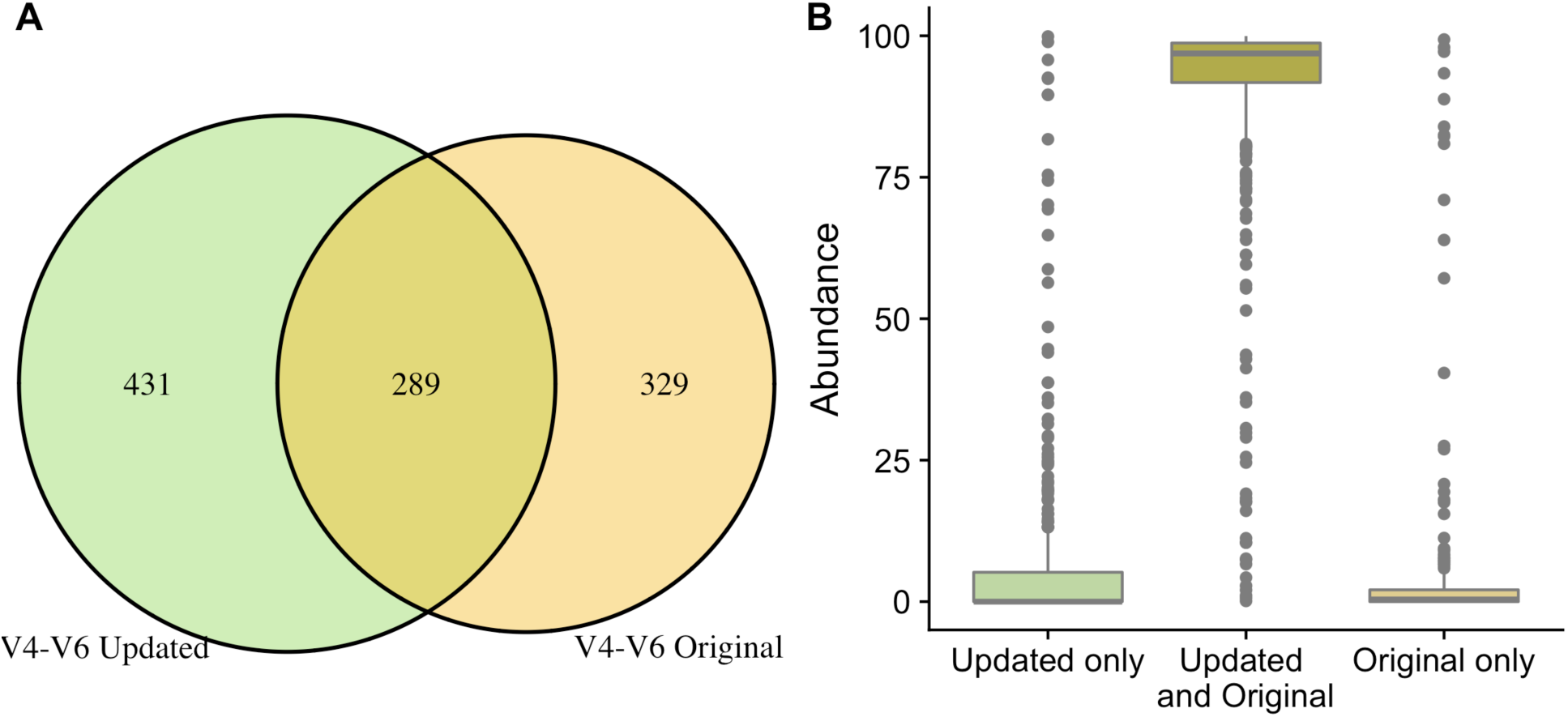
Comparison of taxa recovered through original and updated processing. OTU tables from the original study were obtained and compared to data generated from ASV tables after repeat processing. Figure 3A depicts the number of individual genera that were identified after the original and updated processing pipelines. There were 329 and 432 unique genera identified with the original and updated pipelines, respectively. A total of 289 genera were identified with both processing pipelines. Figure 3B shows the relative sequence abundances of those identified with only original or updated processing, as well as those identified with both pipelines. As depicted, the unique genera that are only identified in the original or updated pipelines tend to be low abundance sequences, while the large majority of the highly abundant sequences were identified through both processing pipelines.

### Amplicon Comparison

A total of 164/207 (79%) of samples had paired V1-V3 and V4-V6 data. Of these, 113 genera were only represented in the V1-V3 dataset, 279 were only represented in the V4-V6 dataset, and 420 were overlapping and represented in both the V1-V3 and V4-V6 datasets (Figure 4a). Like patterns detected when comparing OTU and ASV tables, the most abundant genera were overlapping and identified in both amplicons, while non-overlapping genera were identified in low abundance sequences (Figure 4b). Of the taxa that were represented in both V1-V3 and V4-V6 amplicons, the median abundance was 99.3%. Of the taxa that were only represented in V1-V3, the median abundance was 0.35%; of the taxa that were only represented in V4-V6, the median abundance was 0.88%. Taxa recovered per sample from V1-V3 and V4-V6 regions are shown in Figure 5. Paired abundances from V1-V3 and V4-V6 regions from the most highly abundant genera are summarized in Figure 6, with the remaining genera summarized in Supplemental Figure 2. A higher relative abundance of *Lactobacillus* was identified with the V1-V3 amplicon, while slightly higher relative abundances of *Gardnerella, Tepidomonas, Escherichia/Shigella*, and *Acidovorax* were identified with the V4-V6 amplicon, with subtle differences in other genera. Without further testing and validation, it is unknown which of these two amplicons are more accurate. However, multiple factors led us to infer that the V4-V6 data might be more reliable in this dataset. First, in earlier stages of processing, it was noted that the V1-V3 amplicon contained many sequences mapping to non-bacterial taxa (e.g., archaea, eukaryote, or not assigned) when compared to the SILVA reference database while V4-V6 amplicon data mapped mainly to bacterial taxa, as expected. Secondly, the V1-V3 amplicon recovered *Gardnerella* in a sparser manner than the V4-V6 amplicon. *Gardnerella* are biologically expected when reviewing prior urinary and vaginal microbiome data. Based on these considerations, we considered the V4-V6 amplicon data to be more reliable, and data from this amplicon were selected for statistical analyses. This mirrors the original analysis, in which the authors elected to focus only on V4-V6 region sequencing results in their publication.(5)

**Figure 4:**
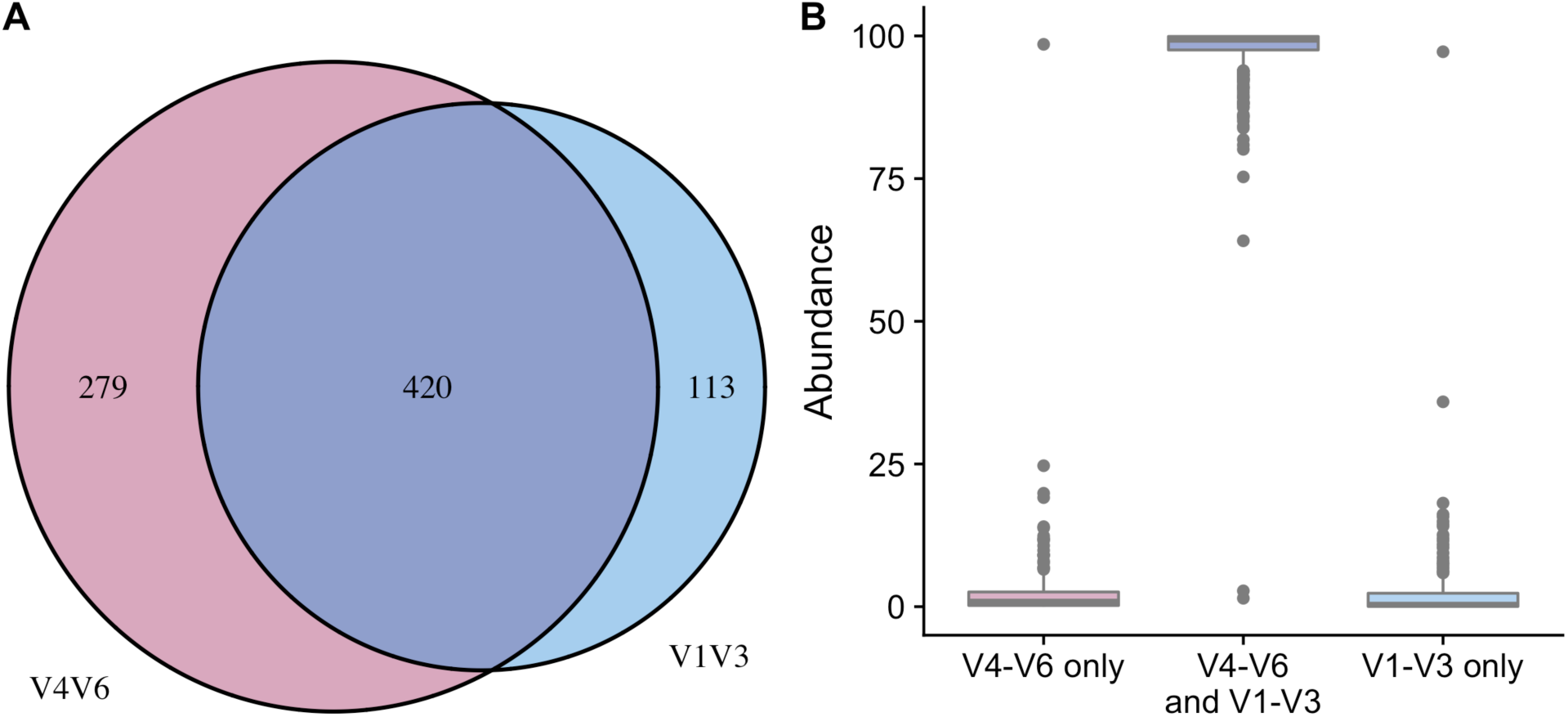
Comparison of taxa recovered from different sequencing amplicons. ASV tables generated through updated processing of V1-V3 and V4-V6 amplicon sequencing data with a DADA2 pipeline were compared. Figure 4A shows that 113 unique genera were identified with the V1-V3 amplicon, 279 unique genera were identified with the V4-V6 amplicon, while 420 genera were shared and identified with both amplicons. Figure 4B shows the relative sequence abundances of those identified with each region, as well as those that were identified with both regions. The unique genera that are only identified with one amplicon are extremely low abundance sequences with an occasional outlier, while the large majority of the highly abundant sequences were identified with both amplicons (i.e., V1-V3 and V4-V6 regions).

**Figure 5:**
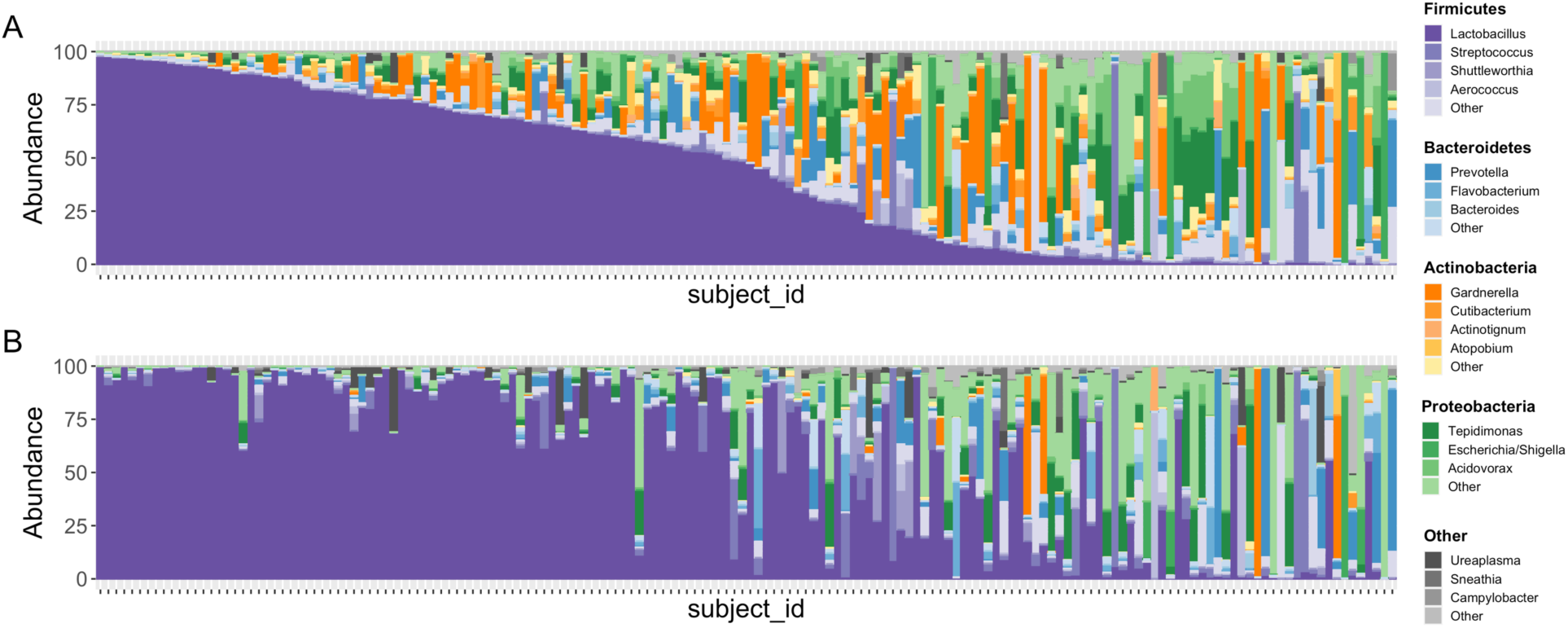
Stacked bar plots illustrating relative abundances of taxa in 167 samples with paired V1-V3 and V4-V6 data. Figure 5A depicts the taxa recovered with the V4-V6 amplicon while Figure 5B depicts the taxa recovered with the V1-V3 amplicon. Each vertical bar depicts an individual sample with plots aligned to compare recovery of data from the same sample in each amplicon. Phyla are assigned distinct colors (e.g., Firmicutes = purple, Bacteroidetes = blue, Actinobacteria = orange, Proteobacteria = green) with individual genera shaded differently. The most intense color shade within each phylum refers to the most abundant genus identified. Though many genera are recovered in similar abundances between both amplicons, *Gardnerella* is one that is noticeably different, with substantially more identified in sequencing data generated from the V4-V6 region.

**Figure 6:**
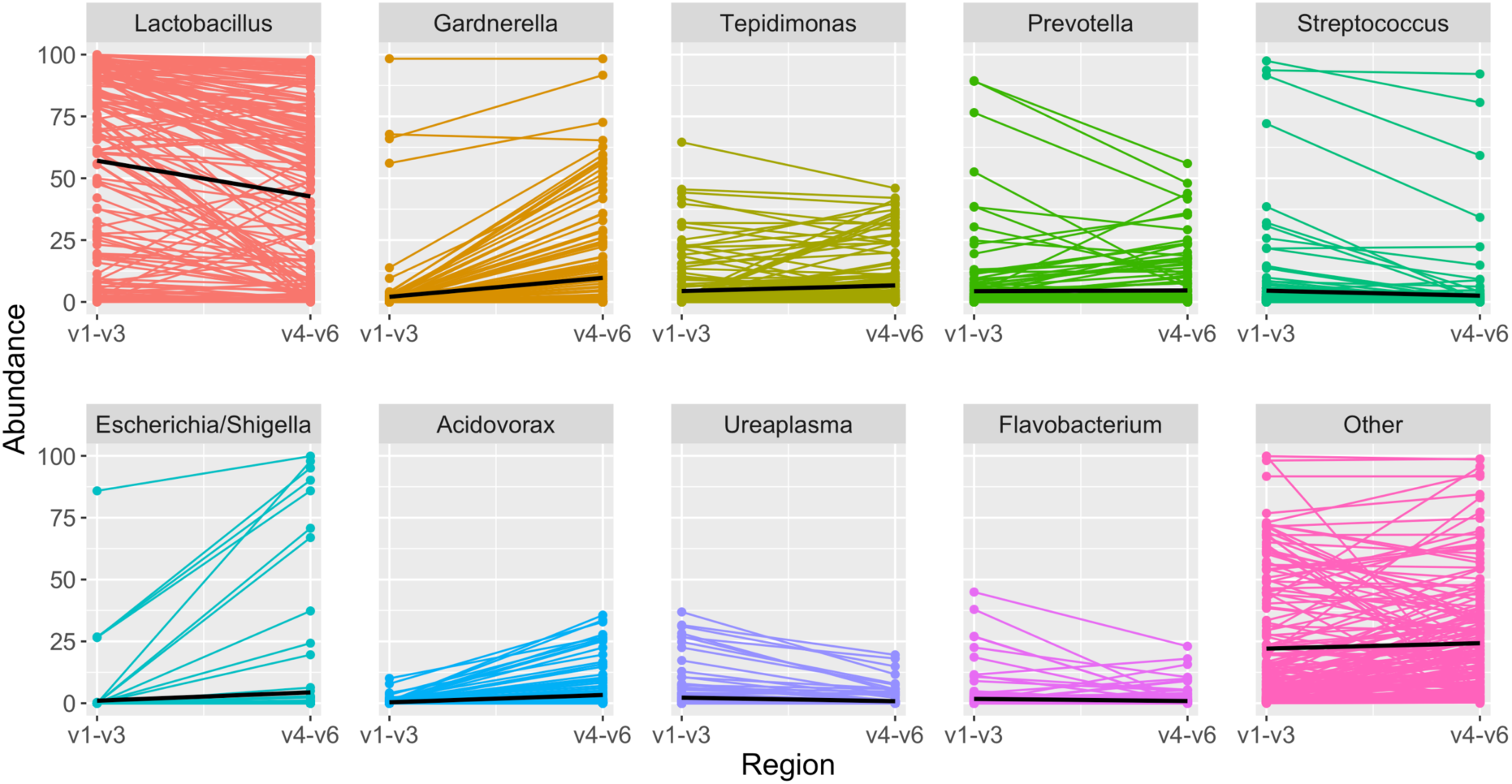
Highest abundance genera among paired samples. Each panel depicts the relative abundance of one genus. On the left is the relative abundance from the V1-V3 amplicon, connected by a line to the right, which shows the relative abundance in the same sample when identified from the V4-V6 amplicon. In each panel the black line summarizes the median abundances across all paired samples. When comparing results from the same sample sequenced with two different amplicons, *Lactobacillus* was identified in slightly higher abundance with the V1-V3 amplicon, while other genera including *Gardnerella, Tepidomonas, Escherichia/Shigella*, and *Acidovorax* were identified in slightly higher abundance with the V4-V6 amplicon.

### Rare ASVs: Filtering Thresholds

After initial review, the microbial data contained within this dataset were mainly dominated by a few ASVs. We filtered data based on read counts, given the low biomass sample type and potential concerns about contamination. We assessed multiple filtering levels ranging from 0.05 - 0.00001 and the effects on downstream clustering. Coarse filtering thresholds of 0.05 – 0.001 resulted in substantial data loss that significantly affected clusters. Once filters less than 0.001 were applied, there were only small changes in clustering results suggesting any of these thresholds could be reasonable. We tested the smaller thresholds of 0.00005 and 0.00001 (the latter removing any sequences where counts were <0.00001 and thus keeping the most data) in clustering. In general, for both DMM and DTMM methods, the filtering levels (less than 0.001) slightly affects the constituents (DMM) and/or number of clusters (DTMM) but had minimal impact on final results after multivariable modeling.

### Clustering into Microbial Communities

As was done in the original analysis, we clustered microbiome data into microbial communities using DMM modeling. In DMM modeling, the number of final clusters are pre-specified. Since the original analysis selected 6 clusters, we chose the same number for the updated analyses. Figure 7 shows the 6 DMM clusters (i.e., microbial communities) that we identified with reprocessed data grouped by MUI and control phenotypes. We also clustered using DTMM modeling where the number of clusters are mathematically chosen based on the data. Using the DTMM approach, there were only 3 clusters when filtering at 0.00005, though a 4^th^ cluster appeared when using a less stringent filtering threshold of 0.00001 (Figure 8). In sensitivity analyses, we evaluated how clustering methodology (DMM vs. DTMM) and filtering thresholds (0.00005 vs. 0.00001) affected associations between microbial community types and MUI versus control status, while controlling for covariates in a multivariable model.

**Figure 7:**
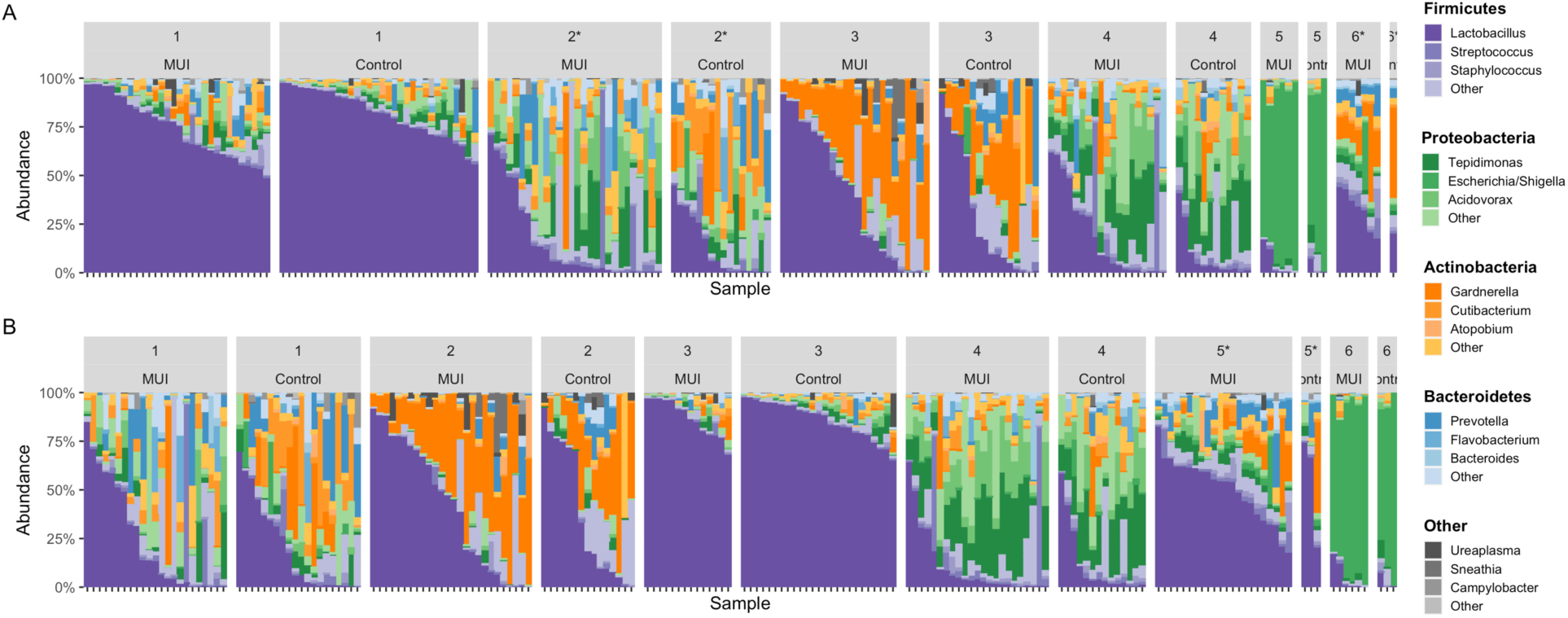
Stacked bar plots illustrating results when combining ASVs from individual samples into microbial communities using DMM clustering. For DMM clustering a total of 6 clusters were chosen *a priori*. Clusters are numbered with each cluster further organized by those samples originating from women with MUI versus control. Figure 7A shows DMM clustering results with the least restrictive filtering threshold of 0.00001, where downstream statistical analyses also identified significant associations between Cluster 2 and Cluster 6 with MUI status. Both Clusters 2 & 6 include low abundances of *Lactobacillus* and a mixture of other genera. Figure 7B shows DMM clustering results with a more restrictive filtering threshold of 0.00005. In this analysis, Cluster 5 (which contains moderate abundance of *Lactobacillus*) is associated with MUI while there was a trend towards Cluster 3 (containing high abundance *Lactobacillus*) being associated with controls.

**Figure 8:**
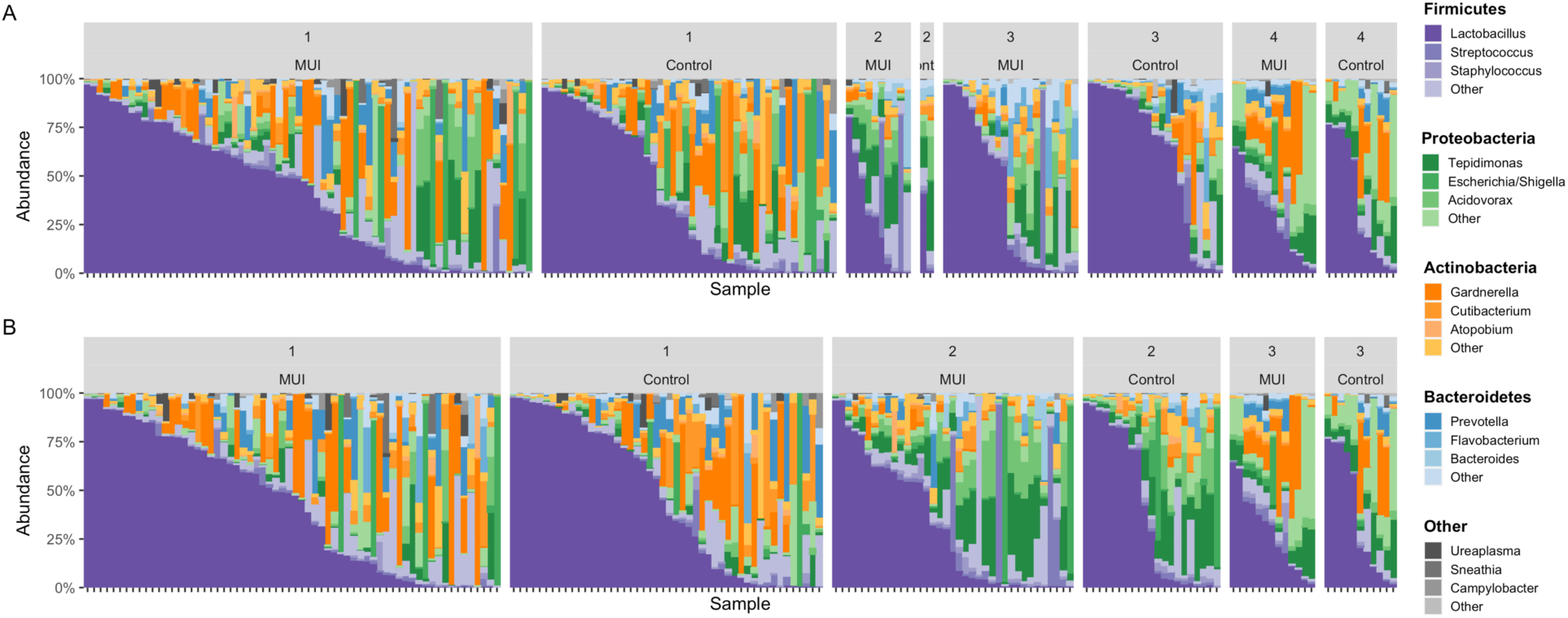
Stacked bar plots illustrating results when combining ASVs into microbial communities using DTMM clustering. For DTMM clustering, the total number of clusters are mathematically chosen during analysis. Clusters are numbered with each cluster further organized by those samples originating from women with MUI versus control. Figure 8A shows DTMM clustering results with the least restrictive filtering threshold of 0.00001, where 4 total clusters were identified. Figure 8B shows DTMM clustering results with a more restrictive filtering threshold of 0.00005, where 3 total clusters were identified. Regardless of the filtering threshold, microbial communities identified through DTMM clustering were not associated with MUI versus control status.

### Testing for Associations between Microbial Communities and MUI versus Control Phenotype

Multivariable models were used to determine whether microbial communities were associated with MUI versus control status, while controlling for other relevant covariates. We first created models that incorporated the clinical site. While some of the sites were significantly associated with outcomes, associations with clinical site were not stable among different models (see supplemental information). To avoid overfitting models, clinical site was removed in final models, which incorporated microbial communities and the following covariates: age, smoking status, ethnicity, BMI, composite menopausal/hormonal status, vaginal pH, history of recurrent UTI, and number days from the most recent catheterization.

The updated analyses that best approximates what was performed in the original analyses was one where DMM methodology was used to cluster samples into microbial communities, and the least stringent filtering threshold (0.00001) was used (Table 2). In this model, Cluster 2 (p < 0.05) and Cluster 6 (p = 0.01) were significantly associated with MUI while controlling for other covariates, including those that were also significantly associated with the MUI outcome, such as BMI (p < 0.01) and Latina ethnicity (p=0.02). Cluster 4 (p=0.09) also trended towards an association with MUI outcomes. In review of the actual taxa within clusters, it appears that the reference group in this model (Cluster 1) was characterized by very high abundances of *Lactobacilli* (see Fig. 7) and was the only group with a higher number of controls compared to MUI, despite the fact that control samples were under-represented in the overall dataset (∼40% of overall samples). A general inference is that communities with high proportions of *Lactobacilli* are associated with control status and communities with lower *Lactobacilli* and higher relative proportions of a <u>combination</u> of Proteobacteria and Actinobacteria are associated with MUI status. Contrary to the original analysis, we did not find that results differ based on age, perhaps due to the inclusion of other covariates such as the composite menopausal/hormonal status or history of recurrent UTIs.

**Table 2:**
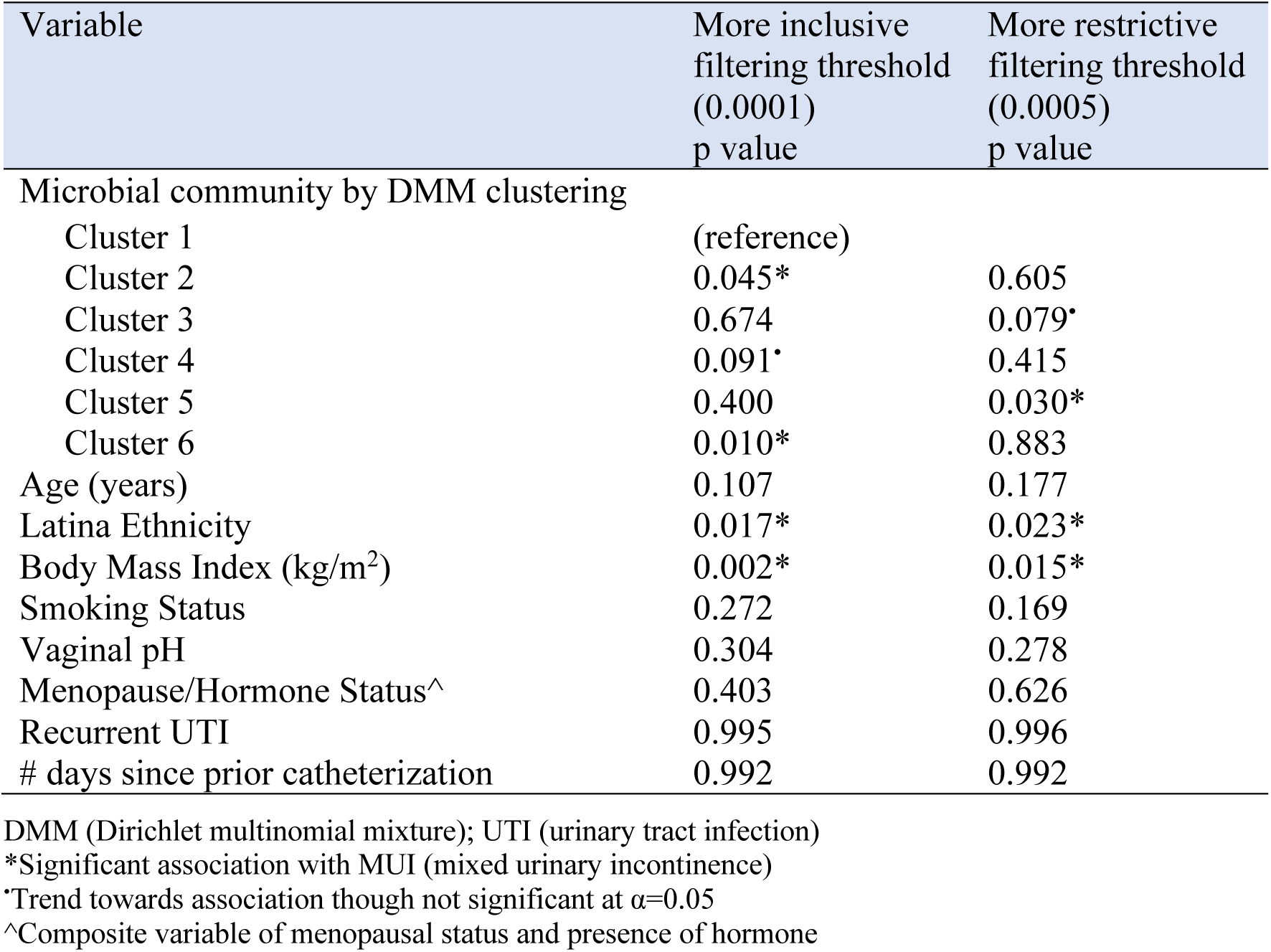
Updated analysis multivariable model testing for associations between MUI versus control

We also performed multiple sensitivity analyses where we used the same modeling approach but with different filtering thresholds and different clustering methodologies. Results are displayed in Table 2 and Supplemental Figure 1. In the model with DMM clustering methodology and a more stringent filtering threshold (0.00005), Cluster 5 was associated with MUI (p=0.03) with a trend towards cluster 3 being associated with controls (p=0.08). Cluster 5 refers to one with moderate abundance of *Lactobacilli*, with some representation of Proteobacteria, Actinobacteria, and Bacteroidetes (see Fig. 8). Cluster 3 has much higher abundance of *Lactobacilli*, with very small components of the others. The covariates BMI and Latina ethnicity remained significantly associated with MUI, even when controlling for other variables, including microbial community types. In essence, this model still shows that some microbial communities are associated with MUI, but the clinical inferences are more challenging to discern.

When incorporating microbial communities generated from DTMM clustering, a more stringent filtering threshold (0.00005) resulted in 3 clusters while a less stringent threshold of 0.00001 resulted in 4 clusters. Regardless of which filtering threshold or number of clusters, there were no associations between microbial communities and MUI outcomes. In these models, BMI and Latina ethnicity still remained significantly associated with MUI (p=0.01, p=0.004, respectively for model with 0.00001 filtering threshold; p=0.007, p=0.004, respectively for model with 0.00005 filtering threshold).

## Discussion

Overall, updated bioinformatic and statistical analyses did not substantially alter the major results of a prior urinary microbiome study. However, this updated analysis offered some interesting nuances that may enhance clinical inferences. We found that an updated bioinformatic processing pipeline recovers many different taxa compared to prior bioinformatic techniques, though most of these differences exist in low abundance taxa that occupy a small proportion of the overall microbiome. We also found that the region of the 16S rRNA gene that is chosen for sequencing can impact downstream results. For the most common (highest abundance) taxa, information will be recovered regardless of the bioinformatic strategy.

However, less abundant taxa may have different biases based on the bioinformatics and sequencing amplicon chosen. For less abundant taxa, results may require additional validation and should be considered carefully when attempting to make inferences. For downstream analyses after bioinformatic processing, the clustering methodology and filtering thresholds that are chosen may affect interpretation of overall results.

Strengths of our approach include the application of techniques that improve precision when analyzing low biomass samples. The bioinformatic processing pipeline applied in this study (i.e., DADA2) corrects for sequencing errors and chimeric sequences to improve accuracy. For updated processing we also used a different reference database (i.e., SILVA), since Greengenes, a database used in many prior urinary microbiome studies, has since been shown to have poor representation of bladder bacteria (6) and has not been updated since 2013. However, our bioinformatic approach is limited as specific expertise (e.g., knowledge of how to use R and other microbiome processing software like QIIME2^7^) may be required compared to prior “plug and play” approaches like the Illumina BaseSpace software. With enhancements in precision, we also encountered more data loss, as some samples did not have high enough quality sequencing information to provide taxonomic data. While we acknowledge that this may decrease the sample size, it may inherently be more scientifically rigorous to remove lower quality sequencing information. Despite technical differences in how sequencing data are handled, our updated processing identified a similar number of phyla and classes compared to the originally processed data (Table 1), with significantly more orders, families, and genera compared what was originally reported.

Along with evaluating the bioinformatic processing techniques, we also tested multiple aspects of statistical analyses, including various filtering and clustering approaches. Generally, investigators need to decide if they want to filter at a lower threshold, thereby keeping more sequencing data. With this approach, there is a risk of over-interpreting data in low biomass samples based on possible contaminants or low abundance sequence information. The other alternative is to filter at a higher threshold, which removes more data, but could result in missing an important association because clusters are less refined. This concept is illustrated in our study when evaluating multivariable models using DMM clustering to create microbial communities. In models with the least stringent filtering threshold, there were associations that appeared both statistically and biologically meaningful. When using the same clustering methodology with a more stringent filtering threshold, there are still statistically significant associations, but the clusters are less refined, and the biologic inference is more obscured. Ultimately, it is only with repeated experiments and ongoing validation that we will expect to understand which approach best approximates the truth. However, it is important for investigators to understand how these choices that are made during statistical analyses may affect downstream results. In future studies, along with good laboratory technique, additional efforts to remove contaminant ASVs during bioinformatic processing (19, 20) may facilitate the ability to use less stringent filtering thresholds since most contaminants will have already been removed.

Existing groups are applying published techniques extrapolated from linear mathematical modeling to analyze microbial datasets. However, many of these techniques contain underlying assumptions of normally distributed data. High dimensional microbial datasets that are used in community-based analyses fail to meet these underlying assumptions, and thus additional techniques are being evaluated and developed. We had hypothesized that tree-based clustering approaches (e.g., DTMM) may be able to better resolve true signal from noise within a dataset. Compared to DMM clustering, DTMM puts more emphasis on lower abundance taxa when clustering. Incorporating a nonparametric mixture as in the available implementation of DTMM also avoids the often-difficult task of pre-specifying the number of clusters. While it was not the case that DTMM clustering was able to better resolve signal from noise in this analysis, it is still possible that other models more akin to machine learning may be useful in the future. For urinary microbiome data it is also not clear if the ratio of high to low abundance taxa (e.g., ratio of *Lactobacilli* compared to other Gram negative & anaerobic bacteria) is more biologically important or if individual low abundance taxa may be important. If the ratio of high abundance bacteria compared to all other bacteria is actually the most biologically important factor, then a clustering method such as DMM that emphasize the highest abundance taxa may actually be preferred.

Compared with the original analysis, we came to slightly different conclusions when evaluating results from our final multivariable models. While we agreed that there were associations between microbial communities and MUI, the context of these associations was different in our updated analysis. Specifically, in the original analysis, 17% of women reported their menopausal status as unknown prompting investigators to dichotomize age based on the approximate age of menopause (51 years) and analyze data in those less than 51 and those older than 51 years. With this approach there were different findings in the two sub-populations,(5) which is somewhat difficult to interpret. Furthermore, hormone status (e.g., whether oral or topical/vaginal hormones were used) were not incorporated into multivariable analyses despite differences noted in MUI and control populations. Multiple investigators have demonstrated that menopause and hormonal status affect microbial compositions in the vagina,(21, 22) and we are now learning that these variables are associated with differences in microbial compositions of the bladder as well.(23) As such, the original clinical data were reviewed to assess how these data were obtained. In this process, we discovered that menopausal information was obtained twice, with one group of questions having more reliable response options. Furthermore, two clinicians (NYS and LB) reviewed all age, menopause, and hormone usage information. Using a combination of these responses, we were able to reliably create a composite variable that incorporated menopausal & hormonal information in an accurate manner. In addition to this composite variable, additional variables that could also confound microbial information such as vaginal pH, history of recurrent UTI, and number days from prior catheterization were also incorporated into multivariable models, while they were not previously. With this modeling strategy, we no longer see age as a separate independent factor affecting microbial community types. Regardless of the modeling strategy used, multiple covariates remained associated with the bladder outcome of MUI, highlighting the importance of incorporating covariates into analyses of microbial data.

With the continued evolution of computational techniques, we expect further improvements and guidelines for analyzing microbial datasets. With this updated analysis, we offer additional insights for investigators embarking on urinary microbiome analyses. Specific consideration should be given to the amplicon (i.e., region of 16S rRNA gene) chosen, as this could affect downstream bacterial identifications. We recommend that investigators consider the bioinformatic processing pipeline, as different options affect specificity of taxonomic information. Furthermore, for optimal taxonomic identifications, investigators should consider reference databases that are updated and contain adequate representation of urinary microbiota. Though default filtering thresholds and clustering methodologies exist, these parameters may need to be optimized based on the questions that are being posed in a microbial dataset. Finally, regardless of how analyses are conducted, multivariable analyses that incorporate potentially confounding clinical variables remain extremely important in analyses of microbial datasets.

## Data Availability

Unprocessed sequencing files are publicly shared on the Sequence Read Archive (SRA), Bioproject ID 703967, Accession #: PRJNA703967
De-identified cinical data are housed at: https://dash.nichd.nih.gov/ under the HMS-ESTEEM study.

https://www.ncbi.nlm.nih.gov/bioproject/PRJNA703967/

## Data Sharing

Unprocessed sequencing files are publicly shared on the Sequence Read Archive (SRA), Bioproject ID 703967, Accession #: PRJNA703967

## Acknowledgments

We would like to graciously acknowledge the women who provided clinical samples as part of the HMS-ESTEEM study, the participating sites from the Pelvic Floor Disorders Network (PFDN), as well as Yuko Komesu, Darrell Dinwiddie, and the University of New Mexico Clinical & Translational Science Center where primary sequencing data were generated. We would also like to acknowledge Ben Carper, Carolyn Huitema and Marie Gantz from RTI International (Research Triangle Park, NC), members of the PFDN Data Coordinating Center who assisted with data transfer and public data sharing. Finally, we acknowledge Karstens laboratory members from Oregon Health & Science University for assistance with data processing (Jean-Philippe Gourdine and Alec Barstad) and preparation of figures (Erin Dahl).

## Contributions to the Field

In this re-analysis of previously generated urinary microbiome sequencing data, updated bioinformatic and statistical analyses offered interesting nuances to the inferences made from the data. An updated bioinformatic processing pipeline recovers many different taxa compared to prior bioinformatic techniques, though most of these differences exist in low abundance taxa that occupy a small proportion of the overall microbiome. We also found that the region of the 16S rRNA gene that is chosen for sequencing can impact downstream results. For the most common (highest abundance) taxa, information will be recovered regardless of the bioinformatic strategy. However, less abundant taxa may have different biases based on the bioinformatics and sequencing amplicon chosen. We again found associations between urinary microbial communities in women and the presence of mixed urinary incontinence. However, when incorporating more covariates related to menopausal status and hormone status, age was no longer separately associated with microbial communities. For downstream analyses after bioinformatic processing, the clustering methodology and filtering thresholds that are chosen may affect interpretation of overall results.

## Footnotes

1 https://greengenes.secondgenome.com/

2 https://twbattaglia.gitbooks.io/introduction-to-qiime/content/lefse.html

3 https://pfdnetwork.azurewebsites.net/

4 https://www.arb-silva.de/

5 Dahl, E., Neer, E., and Karstens, L. (2021). microshades: A custom color palette for improving data visualization. Available at: https://karstenslab.github.io/microshades

6 Mao J and Ma L. (2020) Dirichlet-tree multinomial mixtures for clustering microbiome compositions. https://arxiv.org/abs/2008.00400

7 https://qiime2.org/

## Notes

### Competing Interest Statement

The authors have declared no competing interest.

### Funding Statement

NIA R03AG060082

### Author Declarations

Duke University Institutional Review Board approval (Pro #00102155)

