## Supplemental Information for "Updating urinary microbiome analyses to enhance biologic interpretation"

Supplemental Table 1: Methods and variables for multivariable modeling

| | **Original Analysis** | **Updated analysis** |
|---|---|---|
| Variable Selection Method | …"demographic and medical history variables significantly associated with DMM community type in bivariate analysis ($p<0.05$) and variables associated with MUI versus control and other variables of clinical significance" were selected as covariates.(1) | All clinical variables collected in the HMS-ESTEEM study(2, 3) were ranked^ based on *a priori* knowledge of how each might be associated with microbial community type or MUI. Highest tier of ranked clinical variables were incorporated into multivariable models; certain variables without associations were removed to avoid overfitting models (i.e., avoid exceeding 15 degrees of freedom). |
| Clinical Variables | | |
| Age | Included; due to associations between age and DMM communities, reduced models in age <51 and age >51 created | Included; reduced models not necessary as associations no longer remain when incorporating other covariates |
| Race* | Not included | Tier 1 variable evaluated in preliminary models; not included in final model (lack of association) |
| Ethnicity (Latina vs not Latina)* | Included | Included |
| BMI | Included | Included |
| Recurrent UTI (3 or more in prior year)* | Not included despite associations with MUI status in bivariate analysis | Included |
| Smoking Status (active vs not active) | Included | Included |
| Menopausal Status | Not included | Included as composite variable[&] |
| Hormone Status | Not included despite associations with MUI status in bivariate analysis | Included as composite variable[&] |
| Vaginal pH | Not included | Included |
| Number days since prior catheterization | Not included | Included[$] |

| | | |
|---|---|---|
| Technical Variables | | |
| Clinical Site | Model ”included a random effect for clinical site.”(1) | Tier 2 variable included in preliminary models; removed from final model due to inconsistent associations and to avoid overfitting models. |
| Sample processing time | Not included | Not included[#] |

DMM = Dirichlet multinomial mixture, HMS-ESTEEM = Human Microbiome Study in the Effects of Surgical Treatment Enhanced with Exercise for Mixed Urinary Incontinence randomized trial, MUI = mixed urinary incontinence, BMI = body mass index, UTI = urinary tract infection

^Variables ranked into three tiers with Tier 1 comprising 10 variables listed above with highest likelihood for having associations with urinary microbes and/or clinical condition. Tier 2 variables included parity, responses on validated questionnaires, clinical site, and presence of detrusor overactivity on urodynamic testing; Tier 3 variables were all others.

*Based on patient self-report

[&]Menopausal status (e.g., premenopausal, postmenopausal, unknown) was collected but 17% responded as “unknown”. Thus, this variable was not included in initial HMS-ESTEEM analysis. Upon detailed review, clinicians were able to resolve the “unknown” responses by considering 3 variables in context with each other: age, menopausal status, and use of estrogen hormone. Thus, a composite variable was derived that incorporates both menopausal & hormonal status with 3 potential responses: 1) postmenopausal, no hormones; 2) post-menopausal any hormones (oral, transdermal, vaginal); 3) premenopausal.

[$]Number of days since prior catheterization was assessed in preliminary models as both a continuous variable and as a categorical variable based on clinical assessment of the level of risk of modifying urinary microbial communities (<30 days; 30-90 days; >90 days).

[#]8 samples with prolonged processing time qualitatively explored. Samples were processed 4-6 days after collection due to weekend shipping (compared to within 2 days per study protocol). All were maintained and shipped with DNA protectant (Assay Assure™) and no differences were noted in data quality.

A

```
## Coefficients:
##                Estimate Std. Error z value Pr(>|z|)
## (Intercept)     -0.1032     0.4254  -0.243   0.8083
## cluster.dmm2    -0.4277     0.4354  -0.982   0.3259
## cluster.dmm3    -0.3748     0.4406  -0.851   0.3949
## cluster.dmm4    -0.4859     0.4897  -0.992   0.3211
## cluster.dmm5    -0.1658     0.8937  -0.186   0.8528
## cluster.dmm6    -1.9091     1.1541  -1.654   0.0981 .
## SiteCC           0.8860     0.5650   1.568   0.1169
## SiteDUKE         0.3436     0.5483   0.627   0.5309
## SiteKaiser_SD -16.1058   757.1010  -0.021   0.9830
## SitePENN        -0.6191     0.6921  -0.894   0.3711
## SitePITT         0.6096     0.7141   0.854   0.3933
## SiteUAB         -0.3913     0.5332  -0.734   0.4630
## SiteUCSD         0.1743     0.7048   0.247   0.8047
## SiteUNM          0.3455     0.5330   0.648   0.5169
## ---
## Signif. codes:  0 '***' 0.001 '**' 0.01 '*' 0.05 '.' 0.1 '
```

B

```
## Coefficients:
##                             Estimate Std. Error z value Pr(>|z|)
## (Intercept)                -31.81650 3862.77351  -0.008   0.9934
## cluster.dmm2                -1.52415    0.89666  -1.700   0.0892 .
## cluster.dmm3                -0.33684    0.83699  -0.402   0.6874
## cluster.dmm4                -1.48898    1.00830  -1.477   0.1397
## cluster.dmm5                16.01175 2496.53586   0.006   0.9949
## cluster.dmm6                -2.94422    1.43820  -2.047   0.0406 *
## SiteCC                      -1.37597    1.14432  -1.202   0.2292
## SiteDUKE                    -1.40647    1.13087  -1.244   0.2136
## SiteKaiser_SD              -36.45184 6718.62480  -0.005   0.9957
## SitePENN                    -2.56752    1.15080  -2.231   0.0257 *
## SitePITT                     0.04663    1.45816   0.032   0.9745
## SiteUAB                     -0.95973    1.29735  -0.740   0.4594
## SiteUCSD                    15.57166 2762.33773   0.006   0.9955
## SiteUNM                     -0.51256    1.20235  -0.426   0.6699
## age                          0.05585    0.03567   1.566   0.1174
## BMI                         -0.10278    0.04356  -2.359   0.0183 *
## factor(Smoking == 0)TRUE     0.94791    0.67043   1.414   0.1574
## Vaginal_pH                  -0.51922    0.43291  -1.199   0.2304
## Meno_HT                      0.37596    0.42586   0.883   0.3773
## RUTIs                      -36.17528 6014.39483  -0.006   0.9952
## prior.cath(30,90]            0.96092 4836.83523   0.000   0.9998
## prior.cath(90,999]          36.90293 3862.77224   0.010   0.9924
## ethnicity.ishispanic1       -1.94085    1.13544  -1.709   0.0874 .
## ---
## Signif. codes:  0 '***' 0.001 '**' 0.01 '*' 0.05 '.' 0.1 ' ' 1
```

Supplemental Figure 1: Preliminary modeling results testing for associations with mixed urinary incontinence (MUI) versus control status. Models include data from DMM clustering as the microbial communities; filtering threshold of 0.00001. Fig 1A shows output from a preliminary model incorporating microbial communities and clinical site showing no associations with MUI; Fig 1B shows output from another preliminary model incorporating microbial communities, clinical site, and all clinical covariates with one site demonstrating an association with MUI status. The final model with microbial communities and clinical covariates (site removed) is depicted in Table 2. Similar patterns were noted in all models including those with DMM clusters and filtering threshold of 0.00005, as well as those with DTMM clusters at both filtering thresholds.

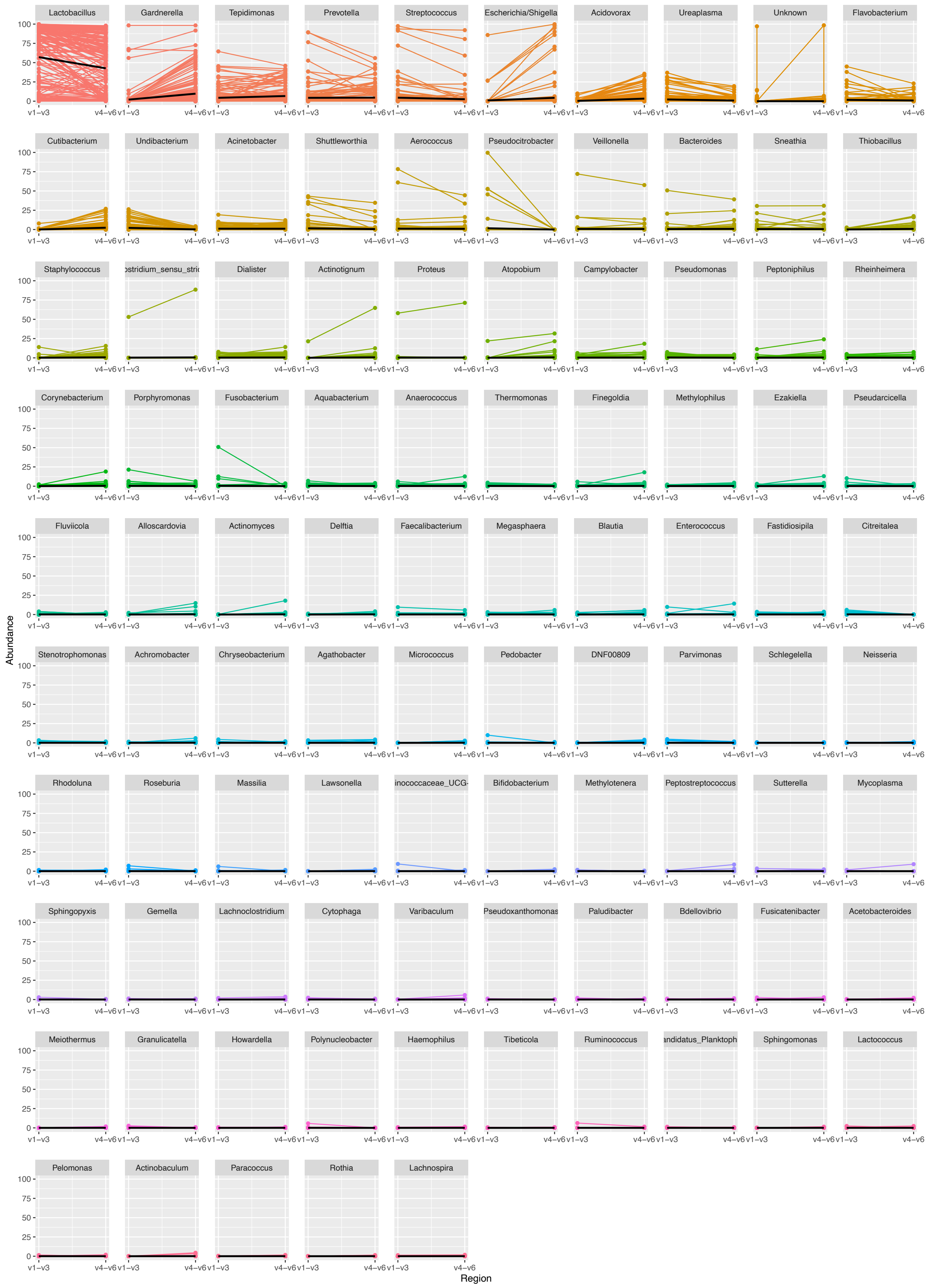

Lactobacillus
Gardnerella
Tepidimonas
Prevotella
Streptococcus
Escherichia/Shigella
Acidovorax
Ureaplasma
Unknown
Flavobacterium
Cutibacterium
Undibacterium
Acinetobacter
Shuttleworthia
Aerococcus
Pseudocitrobacter
Veillonella
Bacteroides
Sneathia
Thiobacillus
Staphylococcus
ostridium_sensu_stric
Dialister
Actinotignum
Proteus
Atopobium
Campylobacter
Pseudomonas
Peptoniphilus
Rheinheimera
Corynebacterium
Porphyromonas
Fusobacterium
Aquabacterium
Anaerococcus
Thermomonas
Finegoldia
Methylophilus
Ezakiella
Pseudarcicella
Fluviicola
Alloscardovia
Actinomyces
Delftia
Faecalibacterium
Megasphaera
Blautia
Enterococcus
Fastidiosipila
Citreitalea
Stenotrophomonas
Achromobacter
Chryseobacterium
Agathobacter
Micrococcus
Pedobacter
DNF00809
Parvimonas
Schlegelella
Neisseria
Rhodoluna
Roseburia
Massilia
Lawsonella
inococcaceae_UCG-
Bifidobacterium
Methylotenera
Peptostreptococcus
Sutterella
Mycoplasma
Sphingopyxis
Gemella
Lachnoclostridium
Cytophaga
Varibaculum
Pseudoxanthomonas
Paludibacter
Bdellovibrio
Fusicatenibacter
Acetobacteroides
Meiothermus
Granulicatella
Howardella
Polynucleobacter
Haemophilus
Tibeticola
Ruminococcus
andidatus_Planktoph
Sphingomonas
Lactococcus
Pelomonas
Actinobaculum
Paracoccus
Rothia
Lachnospira
Abundance
0
25
50
75
100
v1–v3
v4–v6
Region

Supplemental Figure 2: Genera identified among paired samples. Each panel depicts the relative abundance of one genus. On the left is the relative abundance from the V1-V3 amplicon, connected by a line to the right, which shows the relative abundance in the same sample when identified from the V4-V6 amplicon. In each panel the black line summarizes the median abundances across all paired samples. The top 105 (based on abundance) genera are depicted.

### **Supplemental Data - References**

1. Komesu YM, Richter HE, Carper B, Dinwiddie DL, Lukacz ES, Siddiqui NY, et al. The urinary microbiome in women with mixed urinary incontinence compared to similarly aged controls. Int Urogynecol J. 2018;29(12):1785-95.
2. Komesu YM, Richter HE, Dinwiddie DL, Siddiqui NY, Sung VW, Lukacz ES, et al. Methodology for a vaginal and urinary microbiome study in women with mixed urinary incontinence. Int Urogynecol J. 2017;28(5):711-20.
3. Sung VW, Borello-France D, Newman DK, Richter HE, Lukacz ES, Moalli P, et al. Effect of Behavioral and Pelvic Floor Muscle Therapy Combined With Surgery vs Surgery Alone on Incontinence Symptoms Among Women With Mixed Urinary Incontinence: The ESTEEM Randomized Clinical Trial. JAMA. 2019;322(11):1066-76.
